# A pilot surveillance report of SARS-CoV-2 rapid antigen test results among volunteers in Germany, 1^st^ week of July 2022

**DOI:** 10.1101/2022.07.18.22277744

**Authors:** Jannik Stemler, Jon Salmanton-García, Ben Weise, Christina Többen, Carolin Joisten, Julian Fleig, Oliver A. Cornely, the VACCELERATE Consortium

**Author notes:** **Correspondence to:** Jannik Stemler, MD, University of Cologne, Faculty of Medicine and University Hospital Cologne, Translational Research, Cologne Excellence Cluster on Cellular Stress Responses in Aging-Associated Diseases (CECAD), Cologne, Germany, Herderstr. 52-54, 50931 Cologne, Germany.

## Abstract

We hypothesized that reported SARS-CoV-2 infection numbers are underestimated and piloted a point prevalence by rapid antigen testing in the VACCELERATE volunteer registry.

Between July-1 and July-7, 2022, 7/419 (1.67%) tests were positive. Compared to reports of the German Federal Government, our results suggest a 2.39-fold higher prevalence.

Our findings imply that the actual prevalence of SARS-CoV-2 may be higher than detected by current surveillance systems, so that current pandemic surveillance and testing strategies need to be adapted.

## Introduction

With the global surge of Severe Acute Respiratory Syndrome-Coronvavirus-2 (SARS-CoV-2) *Omicron* subvariants BA.4 and BA.5, its enhanced antibody escape, and reduced infection control and prevention measures in most countries, infections due to SARS-CoV-2 rose also among vaccinated and recovered individuals.^1,2^ At the same time, hospitalizations and death rates did not seem to rise significantly, meaning that the infections do not affect most people severely, but still influence people’s daily lives and cause massive work absences.^3^

In Germany, the reported incidence of laboratory-based diagnosis, i.e. primarily via polymerase chain reaction (PCR) of SARS-CoV-2 infections increased rapidly since the beginning of June, 2022 ^4^. Epidemiological predominance of the Omicron BA.5 subvariant was noted from end of the same month.^5^ The numbers reported by the Federal Government are likely to differ substantially from the real incidence as most SARS-CoV-2 infected individuals are not tested via PCR anymore according to the national testing strategy by the Federal Ministry of Health.^6^ They were rather diagnosed by rapid antigen test (RAT), which may not be added to the national statistics.

We aimed to describe the point prevalence of SARS-CoV-2 infections among a representative random sample of volunteers in Germany, to elucidate a potential underreporting in governmental numbers, and to estimate the actual prevalence of SARS-CoV-2 infections.

## Methods and Results

The VACCELERATE Volunteer Registry is an online registry under the umbrella of the VACCELERATE consortium, where people interested in participating in SARS-CoV-2 clinical studies can sign up via online survey (www.vaccelerate.eu/volunteer-registry).^7^ The VACCELERATE Volunteer Registry was approved by the Ethics Committee of the Medical Faculty of the University of Cologne (Cologne, Germany) (Study number 20–1536) and currently is active in 15 European countries.

Among those registered in Germany, 32,962 volunteers ≥12 years were randomly selected with same odds to be shortlisted (0·073%). Selected volunteers were invited to participate in this survey. A sample of 2400 registered individuals ≥12 years of age was contacted via email between June 23^rd^ and 25^th^, 2022 (Figure 1).

**Figure 1.**
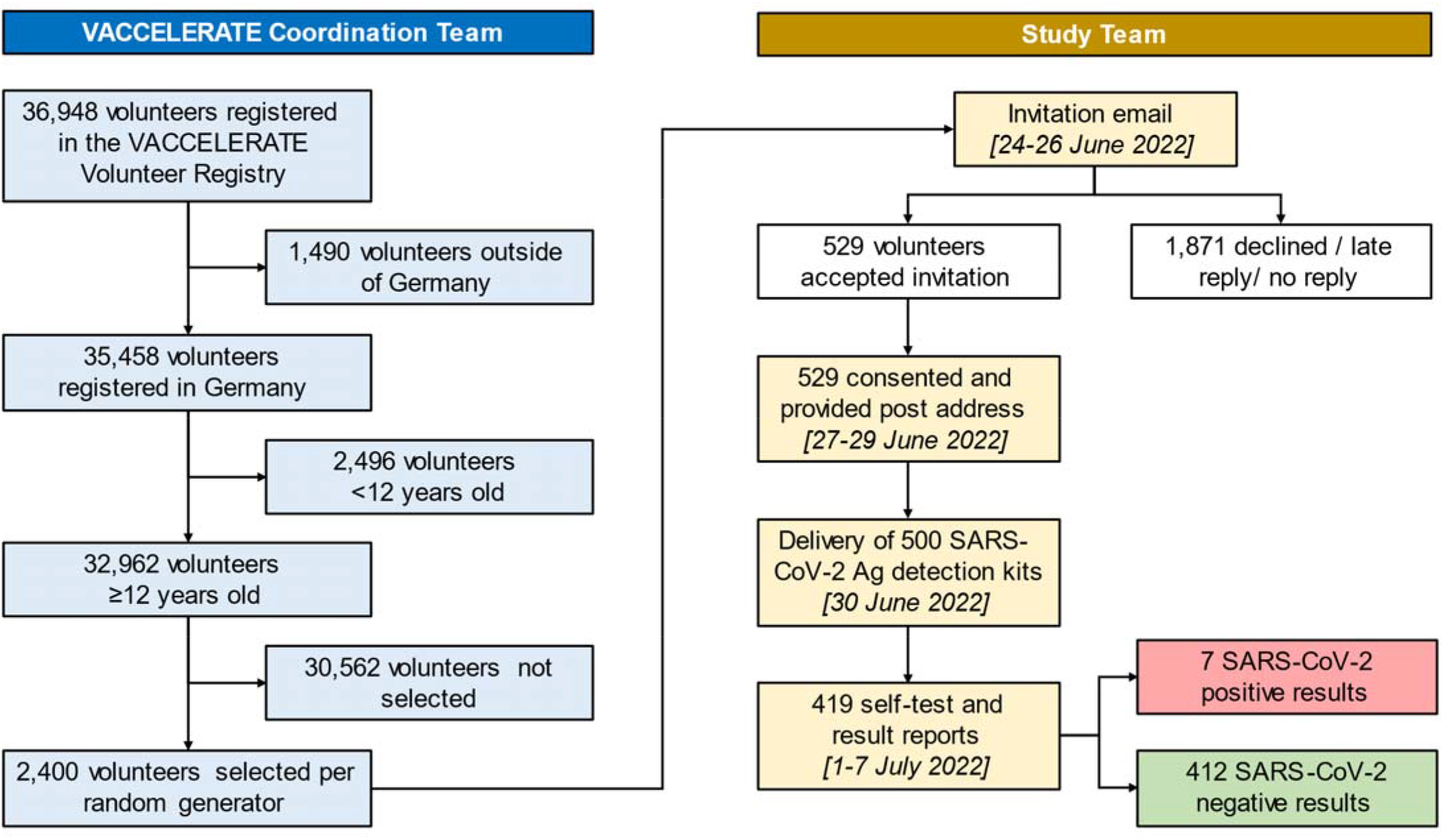
Methodology and enrolment flow chart.

If a volunteer desired to participate, postal address was collected to allow sending of a RAT kit. Then, a SARS-CoV-2 rapid test (“NEWGENE COVID-19 Antigen Test Kit Schnelltest”, COVID-19-NG21, New Gene Bioengineering Co. Ldt., Hangzhou, People’s Republic of China) was sent with instructions on how to self-test properly on the day of delivery. Five-hundred test-kits were sent on June 30^th^, 2022.

Results were then self-reported via email and collected until July 8^th^, 2022 and therefore resemble a point prevalence of July 1^st^ to July 7^th^, 2022. Overall, 58% of participants were female, median age of participants was 44 years (Table 1). Up to this point, 419 participants had submitted a result, among those, seven participants tested positive for SARS-COV-2 via RAT. Six tests were damaged and thus unevaluable or were non-readable (Figure 2).

**Table 1.** Demographic data and test results of participants.

|  | Overall |  | Positive |  | Negative |  |
| --- | --- | --- | --- | --- | --- | --- |
|  | n | % | n | % | n | % |
| **Age, median (IQR) [range]** | 44 (34-57) [12-80] |  | 43 (28-61) [12-65] |  | 44 (34-57) [12-80] |  |
| 12-17 | 11 | 2.6 | 1 | 14.3 | 10 | 2.4 |
| 18-30 | 77 | 18.4 | 1 | 14.3 | 76 | 18.4 |
| 31-40 | 89 | 21.2 | 1 | 14.3 | 88 | 21.4 |
| 41-50 | 74 | 17.7 | 1 | 14.3 | 73 | 17.7 |
| 51-60 | 102 | 24.3 | 1 | 14.3 | 101 | 24.5 |
| 61-70 | 54 | 12.9 | 2 | 28.6 | 52 | 12.6 |
| 71-80 | 12 | 2.9 | 0 | 0.0 | 12 | 2.9 |
| **Gender** |  |  |  |  |  |  |
| Female | 239 | 57.0 | 6 | 85.7 | 233 | 56.6 |
| Male | 180 | 43.0 | 1 | 14.3 | 179 | 43.4 |
| **SARS-CoV-2 rapid antigen test results** |  |  |  |  |  |  |
| Positive | 7 | 1.7 | 7 | 100.0 | - | - |
| *Already known to participant* | 7 | 1.7 | 7 | 100.0 | - | - |
| *Reported to government – yes* | 4 | 1.0 | 4 | 57.1 | - | - |
| *Reported to government – no* | 3 | 0.7 | 3 | 42.8 | - | - |
| Negative | 412 | 98.3 | - | - | 412 | 100.0 |

**Figure 2.**
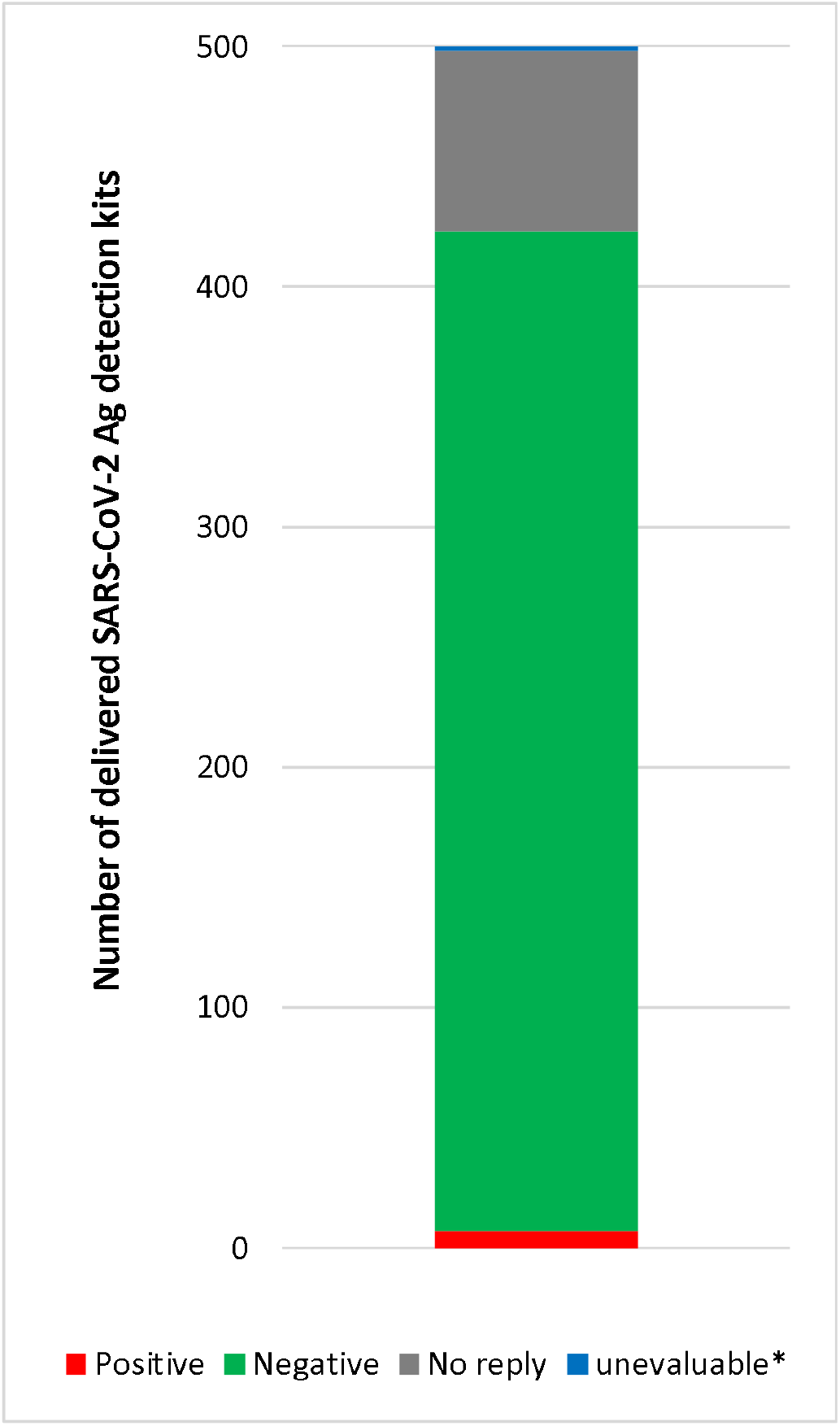
SARS-CoV-2 rapid antigen test (RAT) results in Germany (N=419), July 1^st^ to 4^th^, 2022. *damaged or non-readable RAT

Among the SARS-CoV-2 positive individuals, information was collected on whether infection was already known and whether a PCR result had been performed to calculate any discrepancy between our survey and the governmental reports. All 7 had known about their infection before, 4/7 (57·1%) also had a registered positive PCR result, meaning that 42·8% of participants with a positive RAT were not represented in the federal statistics. Among RAT positive individuals, all 7 had received at least three doses of SARS-CoV-2 vaccine. None of them was hospitalized with COVID-19. Geographic distribution of participants was captured via postal code and showed an even distribution throughout Germany (Figure 3). Evaluation of regional 7-day incidence compared to the weekly governmental report was not meaningful due to small sample number.^8^

**Figure 3.**
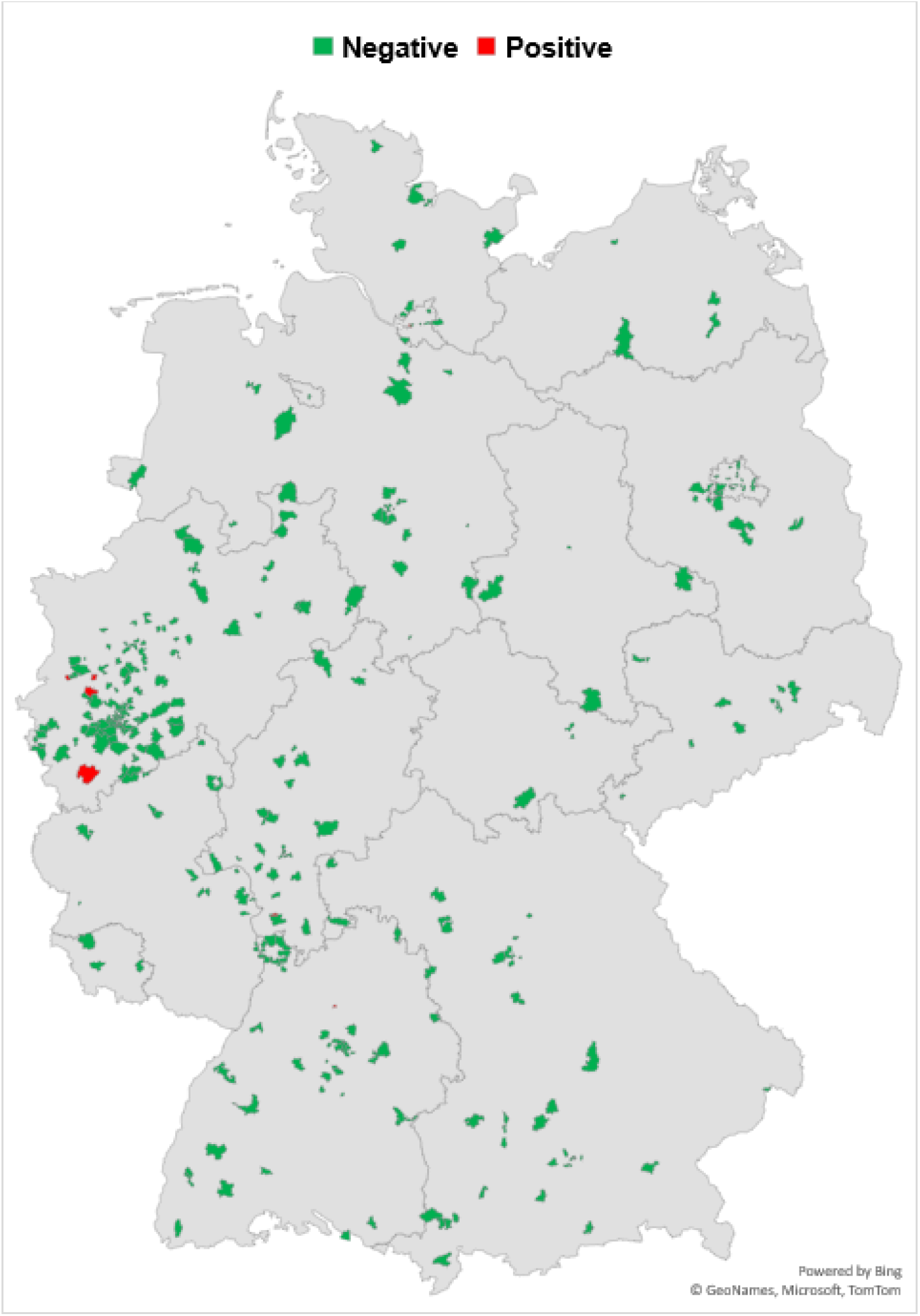
Geographical distribution of test results in Germany, July 1^st^ to 4^th^, 2022.

We report a point prevalence rate for SARS-CoV-2 infection of 1·67% (7/419, equals 1,670 per 100,000 inhabitants) in our dataset. If calculated as a value for a representative random sample of individuals living in Germany, it would describe a 7-day-case number of 1·389·818. This means a 2·39 times higher number than the officially reports the RKI dashboard (https://corona.rki.de/) by July 9^th^, 2022 (7-day incidence of 700,3 and 7-day case number 582,315).

## Discussion and Implications

With increasing SARS-CoV-2 infection numbers among immunized individuals due to the *Omicron* variant, the concurrent antibody-escape of the BA.5 subvariant and without any contact precautions in place, SARS-CoV-2 infections spread even more rapidly than previously observed. This affects large parts of the population with extremely high absolute numbers of simultaneously infected individuals. The current pandemic situation in Germany has affected the so-called critical infrastructure (healthcare, transport sector, industry, agriculture) which is burdened by massive personnel sick leave numbers. Recently, Germany faced noticeable delay of goods and service delivery. At the same time, numbers of hospital admissions and COVID-19 patients in need of intensive care are increasing as of July 8^th^, 2022 in Germany, aggravating the burden on the health care system.^9^

Facing this situation, political action regarding infection control prevention measures seems indispensable. With the third change to the National Testing Strategy of the German Federal Ministry of Health, capacities for SARS-CoV-2 testing were aimed to be utilized more precisely. Therefore, RAT will only be free-of-charge for defined risk groups from July 1^st^, 2022 in Germany.^10^ This may further decrease the reliability of the reported SARS-CoV-2 incidence. Otherwise, the possibility to register self-tested RAT results in the governmental database would allow more precise reporting. Recent regional sewage water sampling of SARS-CoV-2 has suggested a 2-fold higher rate of SARS-CoV-2 infections than the reported incidence,^11^ which is in line with our finding of a 2·39-fold higher prevalence.

Policy makers may use pilot projects like ours to adjust strategies, as well as to guide the local or timely implementation of infection prevention measures. In an expansion of our project, larger and thus more representative groups may be tested regularly in the future in point prevalence studies, while at the same time systematic surveillance of sewage water may be helpful to complement real-time pandemic monitoring.^12^

Our pilot investigation has some limitations. First, we assessed a pilot point prevalence with a rather low participant number. Second, our investigation excluded children below 12 years of age, who may show different epidemiological patterns Third, self-testing bears a potential for inferior test performance.^13^ Fourth, the geographic distribution of reported test results varies substantially and regional epidemiological patterns regarding prevalence of SARS-CoV-2 infections are not displayed.

To conclude, determination of exact numbers of incidence and prevalence in a rapidly globally spreading disease is almost impossible.^14^ Overall, we show that results of random point prevalence studies differ largely from reported governmental data. Such evaluations as well as wastewater analyses may help to determine more precisely a status of the pandemic, when mass testing is not feasible anymore due to capacities or the economic and logistical burden of such testing strategies, or when people infected with SARS-CoV-2 may refrain from confirmatory testing.

## Data Availability

All data produced in the present study are available upon reasonable request to the authors.

## Ethical statement

The VACCELERATE Volunteer Registry was approved by the Ethics Committee of the Medical Faculty of the University of Cologne (Cologne, Germany) (Study number 20–1536).

## Author Contributions

JS: conceptualization, project administration, supervision, methodology, investigation, formal analysis, visualization, writing – original draft, writing – review and editing.

JSG: methodology, resources, investigation, formal analysis, visualization, writing – review and editing.

BW: resources, investigation, formal analysis, writing – review and editing.

CT: investigation, formal analysis, writing – review and editing.

CJ: investigation, formal analysis, writing – review and editing.

JF: investigation, formal analysis, writing – review and editing.

OAC: conceptualization, project administration, resources, supervision, methodology, investigation, formal analysis, writing – review and editing.

## Acknowledgement

We thank Kerstin Albus, Eva Hagemeister, Sarah Heringer, Natalie Irchin, Janina Leckler, Fiona Stewart, and Jan Thielebeule for logistical and administrative support.

